# Artificial intelligence-enabled rapid diagnosis of COVID-19 patients

**DOI:** 10.1101/2020.04.12.20062661

**Authors:** Xueyan Mei, Hao-Chih Lee, Kai-yue Diao, Mingqian Huang, Bin Lin, Chenyu Liu, Zongyu Xie, Yixuan Ma, Philip M. Robson, Michael Chung, Adam Bernheim, Venkatesh Mani, Claudia Calcagno, Kunwei Li, Shaolin Li, Hong Shan, Jian Lv, Tongtong Zhao, Junli Xia, Qihua Long, Sharon Steinberger, Adam Jacobi, Timothy Deyer, Marta Luksza, Fang Liu, Brent P. Little, Zahi A. Fayad, Yang Yang

## Abstract

For diagnosis of COVID-19, a SARS-CoV-2 virus-specific reverse transcriptase polymerase chain reaction (RT-PCR) test is routinely used. However, this test can take up to two days to complete, serial testing may be required to rule out the possibility of false negative results, and there is currently a shortage of RT-PCR test kits, underscoring the urgent need for alternative methods for rapid and accurate diagnosis of COVID-19 patients. Chest computed tomography (CT) is a valuable component in the evaluation of patients with suspected SARS-CoV-2 infection. Nevertheless, CT alone may have limited negative predictive value for ruling out SARS-CoV-2 infection, as some patients may have normal radiologic findings at early stages of the disease. In this study, we used artificial intelligence (AI) algorithms to integrate chest CT findings with clinical symptoms, exposure history, and laboratory testing to rapidly diagnose COVID-19 positive patients. Among a total of 905 patients tested by real-time RT-PCR assay and next-generation sequencing RT-PCR, 419 (46.3%) tested positive for SARSCoV-2. In a test set of 279 patients, the AI system achieved an AUC of 0.92 and had equal sensitivity as compared to a senior thoracic radiologist. The AI system also improved the detection of RT-PCR positive COVID-19 patients who presented with normal CT scans, correctly identifying 17 of 25 (68%) patients, whereas radiologists classified all of these patients as COVID-19 negative. When CT scans and associated clinical history are available, the proposed AI system can help to rapidly diagnose COVID-19 patients.

## Main Manuscript

The coronavirus disease 2019 (COVID-19) pandemic has rapidly propagated due to widespread person-to-person transmission^1–6^. Laboratory confirmation of SARS-CoV-2 is performed with a virus-specific reverse transcriptase polymerase chain reaction (RT-PCR), but the test can take up to two days to complete. Chest computed tomography (CT) is a valuable component of evaluation and diagnosis in symptomatic patients with suspected SARS-CoV-2 infection^4^. Nevertheless, chest CT findings are normal in some patients early in the disease course and therefore chest CT alone has limited negative predictive value to fully exclude infection^7^, highlighting the need to incorporate clinical information in the diagnosis. We propose that artificial intelligence (AI) algorithms may meet this need by integrating chest CT findings with clinical symptoms, exposure history, and laboratory testing in the algorithm. Our proposed joint AI algorithm combining CT images and clinical history achieved an AUC of 0.92 and performed equally well in sensitivity (84.3%) as compared to a senior thoracic radiologist (74.6%) when applied to a test set of 279 cases. While the majority of suspected patients currently have little option but to wait for RT-PCR test results, we propose that an AI algorithm has an important role for the rapid identification of COVID-19 patients which could be helpful in triaging the health system and combating the current pandemic.

The COVID-19 pandemic has resulted in over 3 million cases worldwide. Early recognition of the disease is crucial not only for individual patient care related to rapid implementation of treatment, but also from a larger public health perspective to ensure adequate patient isolation and disease containment. Chest computed tomography (CT) is more sensitive and specific than chest radiography in evaluation of SARS-CoV-2 pneumonia, and there have been cases where CT findings were present prior to clinical symptomatology onset^4^. In the current climate of stress on healthcare resources due to the COVID-19 outbreak, including a shortage of RT-PCR test kits, there is an unmet need for rapid, accurate, unsupervised diagnostic tests for SARS-CoV-2.

Chest CT is a valuable tool for the early diagnosis and triage of patients suspected of SARS-CoV-2 infection. In an effort to control the spread of infection, physicians, epidemiologists, virologists, phylogeneticists, and others are working with public health officials and policymakers to better understand the disease pathogenesis. Early investigations have observed common imaging patterns on chest CT^8,9^. For example, an initial prospective analysis in Wuhan revealed bilateral lung opacities on 40 of 41 (98%) chest CTs in infected patients and described lobular and subsegmental areas of consolidation as the most typical imaging findings^4^. Our initial study with chest CTs in 21 real-time RT-PCR assay confirmed patients also found high rates of ground-glass opacities and consolidation, sometimes with a rounded morphology and peripheral lung distribution^7^. A recent study has also shown that CT may demonstrate lung abnormalities in the setting of a negative RT-PCR test^10^.

During an outbreak of a highly infectious disease with person-to-person transmission, hospitals and physicians may have increased workloads and limited capabilities to triage and hospitalize suspected patients. Previous work demonstrated that early stage coronavirus patients may have negative findings on CT^7^, limiting radiologists’ ability to reliably exclude disease. While waiting 6-48 hours for the confirmation of the SARSCoV-2 coronavirus by RT-PCR, patients who are infected may spread the virus to other patients or caregivers if resources are not available to isolate patients who are only suspected to be infected; nosocomial infection was inferred in approximately 40% of cases in a recent large series^11^. Rapid detection of COVID-19 patients is imperative because an initial false negative could both delay treatment and increase risk of viral transmission to others. In addition, radiologists with expertise in thoracic imaging may not be available at every institution, increasing the need for AI aided detection.

Artificial intelligence (AI) may provide a method to augment early detection of SARSCoV-2 infection. Our goal was to design an AI model that can identify SARS-CoV-2 infection based on the initial chest CT scans and associated clinical information that could rapidly identify COVID-19 (+) patients in the early stage. We collected chest CT scans and corresponding clinical information obtained at patient presentation. Clinical information included travel and exposure history, leukocyte counts (including absolute neutrophil number, percentage neutrophils, absolute lymphocyte number, and percentage lymphocytes), symptomatology (presence of fever, cough, and sputum), patient age, and patient sex (Table 1).

**Table 1.** Characteristics of Patient’s Clinical Information. The two-sided p-clinical feature was tested by logistic regression. The Hosmer-Lemeshow goodness of fit was used to assess the logistic regression fit.

|  | **COVID-19 positive**<br>**(n=419)** | **COVID-19 negative**<br>**(n=486)** | **p-value** |
| --- | --- | --- | --- |
| **Sex** |  |  |  |
| Men | 208 (49.6) | 280 (57.6) | 0.36 |
| <sup>†</sup>Age (years) | 43.0±16.4 (32, 54) | 38.6±16.3 (27, 48) | 0.00086 |
| Temperature (°C) | 37.6±0.9 | 37.5±1.0 | 0.60 |
| Exposure history | 319 (76.1) | 254 (52.3) | <1e-4 |
| **Clinical symptoms** |  |  |  |
| Fever | 314 (75.0) | 318 (65.4) | 0.0089 |
| Cough | 210 (50.1) | 128 (26.3) | <1e-4 |
| Cough with sputum | 83 (20.0) | 177 (36.4) | <1e-4 |
| **Laboratory findings** |  |  |  |
| <sup>†</sup>WBC (10<sup>9</sup>/L) | 5.4±2.2 (4.0, 6.4) | 8.3±3.4 (6.1, 10.0) | 0.025 |
| <sup>†</sup>Neutrophils (10<sup>9</sup>/L) | 3.5±1.9 (2.3, 4.2) | 5.9±3.1 (3.7, 7.4) | 0.11 |
| <sup>†</sup>Percentage Neutrophils | 63.0±14.7 (53.8, 74.2) | 68.9±13.1 (60.7, 78.8) | 0.30 |
| <sup>†</sup>Lymphocytes (10<sup>9</sup>/L) | 1.4±0.8 (0.9, 1.7) | 1.6±0.9 (1.0, 2.0) | 0.12 |
| <sup>†</sup>Percentage Lymphocytes | 26.4±12.5 (17.3, 34.5) | 21.3±11.1 (12.7, 28.9) | 0.53 |

† Data in parenthesis shows Interquartile Range (IQR)
± indicates mean ± standard deviation
Other data in parenthesis shows percentage

We first developed a deep convolutional neural network (CNN) to learn the imaging characteristics of COVID-19 patients on the initial CT scan. We then used support vector machine (SVM), random forest, and multi-layer perceptron (MLP) classifiers to classify the COVID-19 patients according to the clinical information. MLP showed the best performance on the tuning set; only MLP performance is reported hereafter. Finally, we created a neural network model combining the radiologic data and the clinical information to predict COVID-19 status (Fig. 1).

**Fig. 1.**
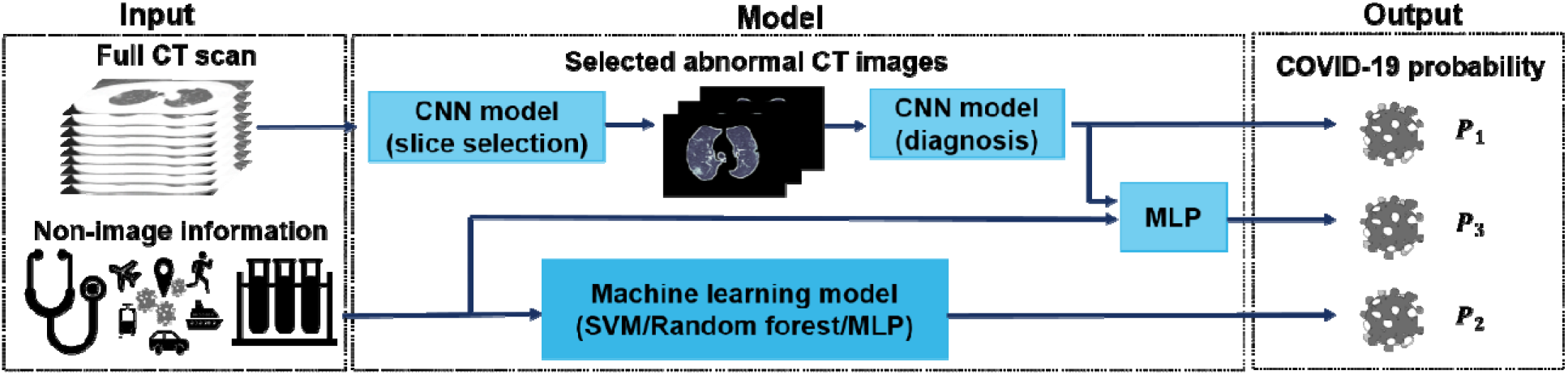
Illustration of the modelling framework. Three AI models are used to generate the probability of a patient being COVID-19 (+): the first is based on a chest CT scan, the second on clinical information; and the third on a combination of the chest CT scan and clinical information. For evaluation of chest CT scans, each slice was first ranked by the probability of containing a parenchymal abnormality, as predicted by the convolutional neural network model (slice selection CNN), which is a pre-trained PTB model that has a 99.4% accuracy to select abnormal lung slices from chest CT scans. The top 10 abnormal CT images per patient were put into the second CNN (diagnosis CNN) to predict the likelihood of COVID-19 positivity (P1). Demographic and clinical data (the patient’s age and sex, exposure history, symptoms and laboratory tests) were put into a machine learning model to classify COVID-19 positivity (P2). Features generated by the diagnosis CNN model and the non-imaging clinical information machine learning model were integrated by a multi-layer perceptron network (MLP) to generate the final output of the joint model (P3). PTB, pulmonary tuberculosis; SVM, support vector machine.

A dataset of the presenting chest CT scans from 905 patients for whom there was a clinical concern of COVID-19 was acquired between Jan 17, 2020 and March 3, 2020 from 18 medical centers in 13 provinces in China. The data set included patients aged from 1 year to 91 years (40.7 year ± 16.5 years), and included 488 men and 417 women. All subjects were acquired using a standard chest CT protocol and were reconstructed using the multiple kernels and displayed with a lung window. A total of 419 patients (46.3%) tested positive for SARS-CoV-2 by laboratory-confirmed real-time RT-PCR assay and next-generation sequencing, while 486 patients (53.7%) tested negative (confirmed by at least two additional negative RT-PCR tests and clinical observation). We randomly split the dataset into 60% training set (534 cases; 242 COVID-19 (+); 292 COVID-19 (-) cases), 10% tuning set (92 cases; 43 COVID-19 (+) cases; 49 COVID-19 (-) cases), 30% test set (279 cases; 134 COVID-19 (+) cases; 145 COVID-19 (-) cases) (Extended Data Fig. 1).

We evaluated the AI models on the test set and compared their performance to one fellowship trained thoracic radiologist with ten years of experience (A. J.) and one thoracic radiology fellow (S. S.). The same initial chest CT and clinical information were available to the radiologists as was provided to the AI model. Sensitivity, specificity and AUC were calculated for both human readers and the AI models. The performance of the AI model and human readers are demonstrated in Fig. 2 and Extended Data Fig. 2. The receiver operating characteristic (ROC) curve of the AI model is shown in Fig. 2.

**Fig. 2.**
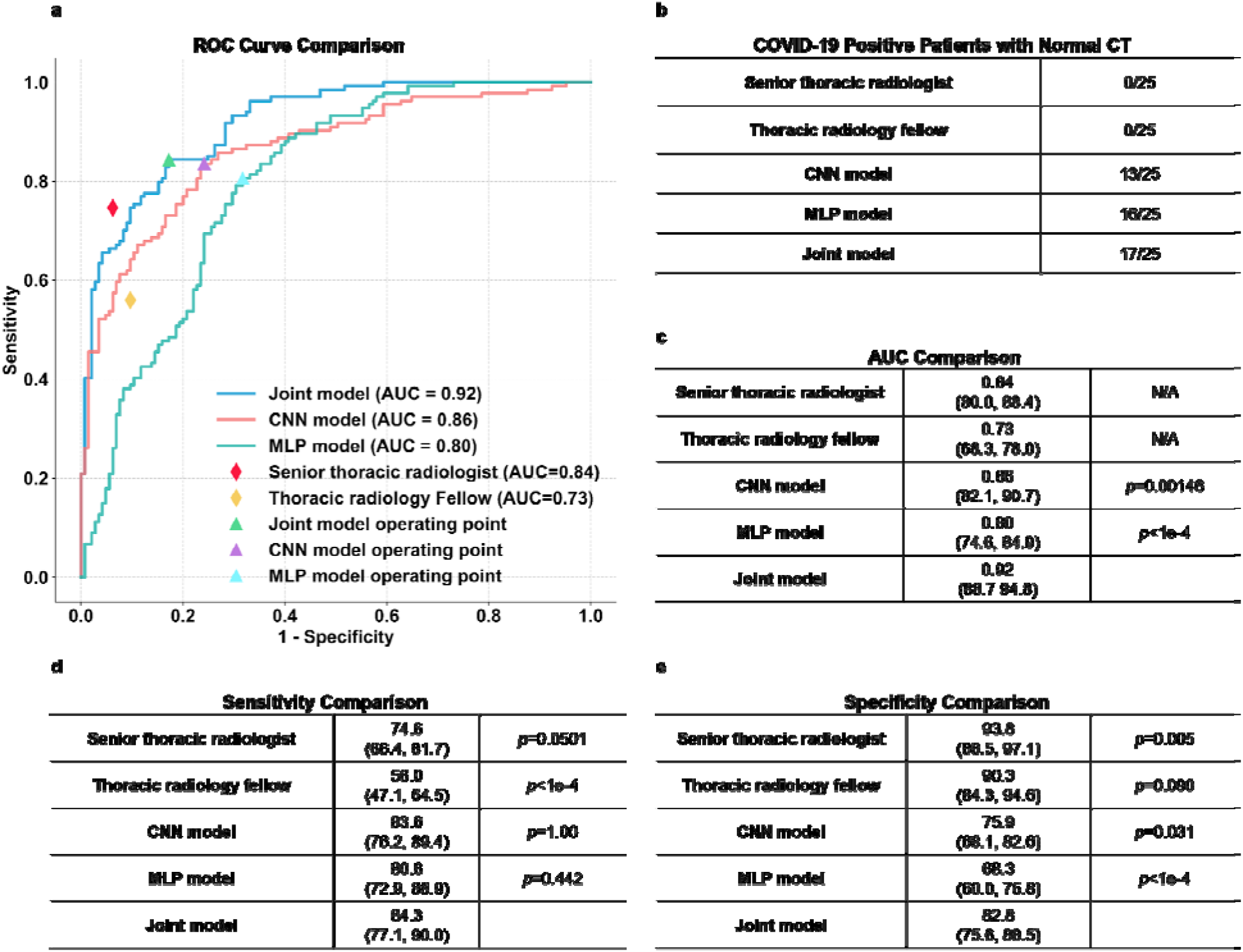
Results of the AI models on the test set. a, Comparison of the ROC curves for the joint model, the CNN model trained based on CT images, the MLP model trained based on clinical information and two radiologists. b, Comparison of the success rates of diagnosing COVID-19 positive patients with normal CT scans. Radiologists were provided with both CT imaging and clinical information in making their diagnoses. c-e, Comparison of the AUCs (c), sensitivities (d) and specificities (e) achieved by the AI models and radiologists. Two-sided p values were calculated by comparing the joint model to the CNN model, the MLP model and the two human readers in sensitivity, specificity and AUC. AUC comparisons were evaluated by the DeLong test; sensitivity and specificity comparisons were calculated by using the exact Clopper-Pearson method to compute the 95% confidence interval shown in parenthesis and exact McNemar’s test to calculate the p-value.

Patient’s age, presence of exposure to SARS-CoV-2, presence of fever, cough and cough with sputum and white blood cell counts were significant features associated with SARS-CoV-2 status. The logistic regression was a good fit (p=0.662). The joint model using both clinical data and CT imaging achieved an 84.3% sensitivity (95% confidence interval (CI) 77.1%, 90.0%), an 82.8% specificity (95% CI 75.6%, 88.5%) and 0.92 AUC (95% CI 0.887, 0.948). The CNN mode that uses only CT imaging data had an 83.6% sensitivity (95% CI 76.2%, 89.4%; *p*=1), a 75.9% specificity (95% CI 68.1%, 82.6%; *p*=0.031) and 0.86 AUC (95% CI 0.821, 0.907; p=0.00146). The multi-layer perceptron (MLP) model that uses only clinical data had an 80.6% sensitivity (95% CI 72.9%, 86.9%; *p*=0.442) and a 68.3% specificity (95% CI 60.0%, 75.8%; *p*<1e-4) and 0.80 AUC (95% CI 0.746, 0.849; *p*<1e-4). The senior thoracic radiologist using both the CT and clinical data achieved a 74.6% sensitivity (95% CI 66.4%, 81.7%; *p*=0.0501), 93.8% specificity (95% CI 88.5%, 97.1%; *p*=0.005) and 0.84 AUC (95% CI 80%, 88.4%). The thoracic radiology fellow using both the CT and clinical data achieved a 56.0% sensitivity (95% CI 47.1%, 64.5%; *p*<1e-4), 90.3% specificity (95% CI 84.3%, 94.6%, *p*=0.090) and 0.73 AUC (95% CI 68.3%, 78.0%). P-value indicates the significance of difference in performance metric compared with respect to the joint model.

With a higher AUC, the joint model integrating CT images and associated clinical information outperformed the model trained on CT images only and the clinical model trained on clinical information only. The joint model, the CT model and the clinical model performed equally well in sensitivity compared to the senior thoracic radiologist but showed statistically significant improvement in sensitivity compared to the thoracic fellow (Fig. 2).

The test set contained 25 COVID-19 (+) patients with a chest CT identified as normal by both of the reading radiologists at presentation. The CNN model identified 13/25 (52%) scans as COVID-19 (+), the clinical model classified 16/25 (64%) as disease positive, and the joint model classified 17/25 (68%) as disease positive, whereas the senior chest radiologist and the chest radiology fellow identified 0/25 (0%) of these scans as disease positive.

We summarized the comparisons of prediction between the joint model and the radiologists in the Extended Data Fig. 3. Of the 134 COVID-19 (+) cases in the test set, 90 out of 134 cases were correctly classified by both the joint model and the senior thoracic radiologist. Forty-four out of 134 cases were classified differently by the joint model and the senior thoracic radiologist. Of the 44 cases, 33 cases were correctly classified positive by the joint model, but were misclassified by the senior thoracic radiologist. Ten cases were classified negative by the joint model, but correctly diagnosed by the senior thoracic radiologist. No cases were misclassified by both the joint model and the senior thoracic radiologist.

**Fig. 3.**
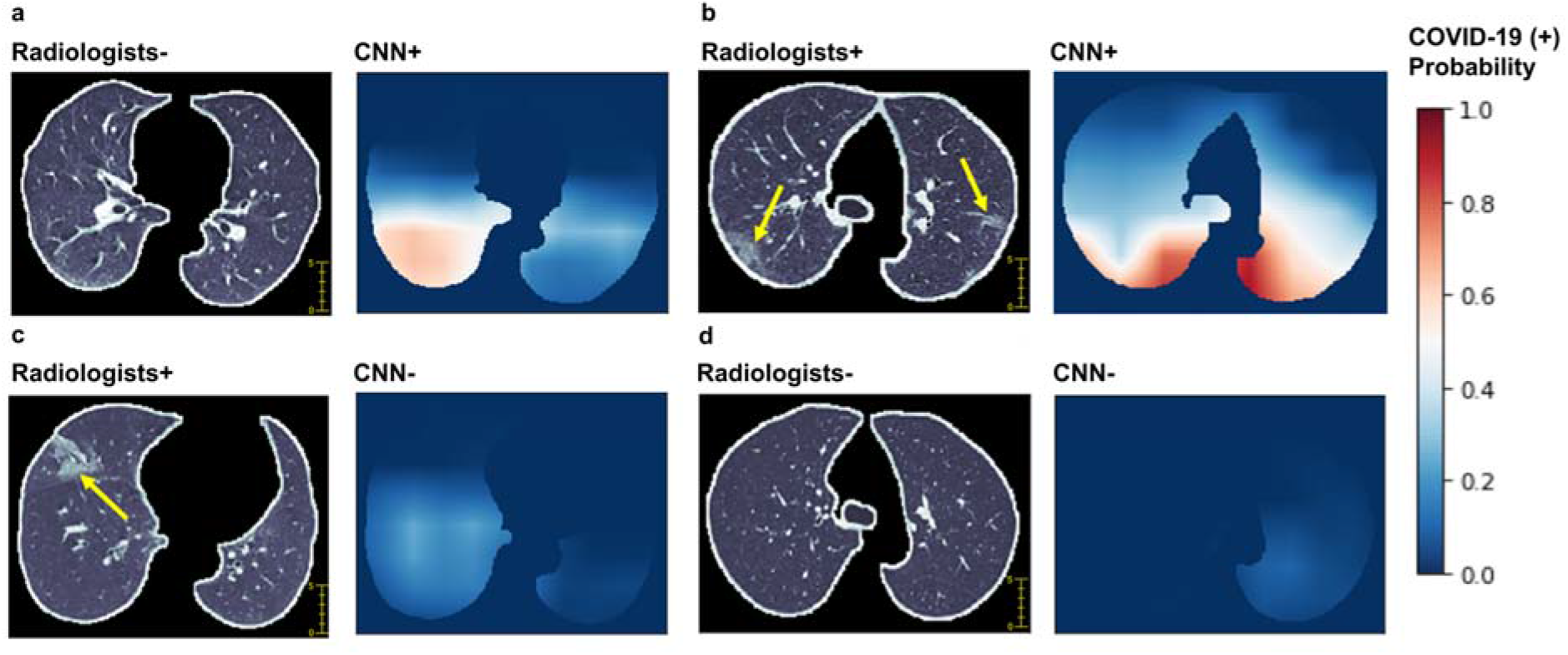
Examples of chest CT images of COVID-19 (+) patients and visualization of features correlated to COVID-19 positivity. For each pair of images, the left image is a CT image showing the segmented lung used as input for the CNN model trained on CT images only, and the right image shows the heatmap of pixels that the CNN model classified as having SARS-CoV-2 affection (red indicates higher probability). (a) A 51-year-old female with fever and history of exposure to SARS-CoV-2. The CNN model identified abnormal features in the right lower lobe (white color), whereas the two radiologists labeled this CT as negative. (b) A 52-year-old female who had a history of exposure to SARS-CoV-2 and presented with fever and productive cough. Bilateral peripheral ground-glass opacities (arrows) were labeled by the radiologists, and the CNN model predicted positivity based on features in matching areas. (c) A 72-year-old female with exposure history to the animal market in Wuhan presented with fever and productive cough. The segmented CT image shows ground-glass opacity in the anterior aspect of the right lung (arrow), whereas the CNN model labeled this CT as negative. (d A 59-year-old female with cough and exposure history. The segmented CT image shows no evidence of pneumonia, and the CNN model also labeled this CT as negative.

Of the 145 COVID-19 (-) cases in the test set, 113 out of 145 cases were correctly classified by both the joint model and the senior thoracic radiologist. Thirty-two out of 145 cases were classified differently by the joint model and the senior thoracic radiologist. Seven cases were correctly classified negative by the joint model, but were diagnosed positive by the senior thoracic radiologist. Twenty-three cases were classified positive by the joint model, but correctly diagnosed negative by the senior thoracic radiologist. Two cases were misclassified by both the joint model and the senior thoracic radiologist.

Chest CT is a well-known diagnostic tool for evaluation of patients with a suspected pulmonary infection. During the outbreak of COVID-19 in some countries including China and South Korea, chest CT has been widely used in clinical practice due to its speed and availability. Most institutions in China have adopted a policy of performing a chest CT scan on any patient with fever and a suspicion of SARS-CoV-2 infection. Initial experience with CT has demonstrated that typical findings are multilobular and bilateral and include both ground glass opacities and consolidation, often with a peripheral lung distribution. Pleural effusions, lymphadenopathy, and discrete pulmonary nodules are very uncommon^9,12,13^. According to the recommendations of the WHO, the most accurate diagnosis of COVID-19 is nucleic acid detection^14^ in secretional fluid collected from a throat swab using RT-PCR. However, there is a shortage of the nucleic acid detection kits and results can take up to two days. Chest CT has also been proposed as an important diagnostic tool. A chest CT study can be obtained and interpreted much more quickly than RT-PCR. While chest CT is not as accurate as RT-PCR in detecting the virus, it may be a useful tool for triage in the period before definitive results are obtained^7,15^. A recent work has implemented AI to differentiate COVID-19 from other pneumonia only based on chest CT images only^16^, which also highlights the necessity for fast and accurate reporting of chest CTs and the potential assistance of AI. In our study, our joint AI model combines CT and clinical data. For patients with mild symptoms demonstrating normal chest CT in the early stage, our model showed that clinical information played a role in the accurate diagnosis of COVID-19.

There are two potential limitations to the use of chest CT. First, the health system during an epidemic may be overburdened, which may limit timely interpretation of the CT by a radiologist. Second, the morphology and severity of pathologic findings on CT is variable. In particular, mild cases may have few if any abnormal findings on chest CT.

We believe implementation of the joint algorithm discussed above could aid in both issues. First, the AI algorithm could evaluate the CT immediately after completion. Secondly, the algorithm outperformed radiologists in identifying COVID-19 (+) patients demonstrating normal CT results in the early stage. Thirdly, the algorithm performed equally well in sensitivity (*p*=0.05) in the diagnosis of COVID-19 as compared to a senior thoracic radiologist. Specifically, the joint algorithm achieved a statistically significant 6% (*p*=0.00146) and 12% (*p*<1e-4) improvement in AUC as compared to the CNN model using only CT images and the MLP model using only clinical information respectively. The AI model could be deployed as an application that can run on a simple workstation alongside the radiologists. Use of the AI tool would require integration with the Radiology PACS and clinical database systems or other image storage database, which is relatively easy to achieve in modern hospital systems. The AI system could be implemented as a rapid diagnostic tool to flag suspected COVID-19 patients when CT images and/or clinical information are available, and radiologists could review these suspected cases identified by AI with a higher priority.

Our proposed model does have some limitations. One major limitation of this study is the limited sample size. Despite the promising results of using the AI model to screen COVID-19 patients, further data collection is required to test the generalizability of the AI model to other patient populations. Collaborative effort in data collection may facilitate improving the AI model. Difficulties on model training also arise due to the limited sample size. In this work we used a pre-trained TB model to select key slices to represent a full 3D CT scan. This approach can reduce computation of training a 3D convolution neural network, with a trade off on missing information in the slices that are not selected for model training and inference. The design of the CNN model offers a natural visualization to explain the prediction. We showcased some examples allowing the AI models to be cross referenced with radiologist’s findings (Fig. 3). However, there are examples in which the visualization fails to provide a clear explanation. We do not know if the model incorporates features such as airways, background of emphysema or the border of the lung in its prediction. Another limitation is the bias towards COVID-19 patients in the training data, which, given the non-specific nature of the ground glass opacity and other features on chest CT images, potentially limits the usefulness of the current AI model to distinguish COVID-19 from other causes of respiratory failure. Therefore, our algorithm may be helpful in places with current high rates of COVID-19 disease, but is unlikely to provide as much usefulness in places or times where COVID-19 prevalence is low. However, the recent study by Li et al demonstrated the ability of AI models to separate COVID-19 from other pneumonias, which is promising for the wider application of this and other AI models.

In future studies, a larger dataset will be collected as the scale of this outbreak is climbing. We aim to explore different approaches in convolutional neural networks including three-dimensional deep learning models and improvement of interpretability of CNN models. The generalizability of the AI system evaluated at multiple centers will be necessary to validate the robustness of the models.

In conclusion, these results illustrate the potential role for a highly accurate AI algorithm for the rapid identification of COVID-19 patients which could be helpful in combating the current disease outbreak. We believe the AI model proposed, that combines CT imaging and clinical information, and shows equivalent accuracy to a senior chest radiologist, could be a useful screening tool to quickly diagnose infectious diseases such as COVID-19 that does not require radiologist-input or physical tests.

## Data Availability

The raw image dataset generated or analysed during the current study are not publicly available due to the dicom metadata containing information that could compromise patient privacy/consent. The models generated by this dataset are publicly available at https://github.com/howchihlee/COVID19_CT.

## Acknowledgements

We thank the computational support by Biomedical Engineering and Imaging Institute at Icahn School of Medicine at Mount Sinai for this work. Z.A.F was funded by US National Institutes of Health grant NIH/NCRR CTSA UL1TR001433 and Mount Sinai COVID Informatics Center. V.M. received funding support from Astra Zeneca, Daiichi Sankyo, Alexion and Horizon Pharma. P.R. was supported by US National Institutes of Health (grant R01-HL071021). Y.Y. was supported by start-up seed fund from Icahn School of Medicine at Mount Sinai. The content is solely the responsibility of the authors and does not necessarily represent the official views of the National Institutes of Health. The funding agencies had no role in study design, data collection and analysis, preparation of the manuscript, or the decision to publish.

## Author Contributions

X.M. and H.L. developed the modeling. X.M., H.L. and C.L. created the figures. X.M., H.L., K.D., T.D., M.H., B.P.L., P.M.R., Z.A.F. and Y.Y. wrote the manuscript. P.M.R., C.C., V.M. and F.L. provided additional guidance in the review process. X.M., H.L., T.D., M.C., A, Z.A.F. and Y.Y. designed the experiments. B.P.L., M.C., A.B., A.J. and S.S. provided clinical expertise. H.S., S.L., K.L., K.D., Q.L., T.Z., B.L., Z.X., J.L., and J.X. collected the dataset. M.L. and F.L. advised on the modeling techniques. A.J. and S.S. evaluated and read the test set cases. X.M. and Y.M. performed statistical analysis. B.P.L., Z.A.F. and Y.Y. supervised the work.

## Methods

### Study participants

The study was approved by the institutional review board of each participating hospital in China and the Icahn School of Medicine at Mount Sinai in New York. The institutional review boards waived the requirement to obtain written informed consent for this retrospective study, which evaluated de-identified data and involved no potential risk to patients. To avert any potential breach of confidentiality, no link between the patients and the researchers was made available.

We collected the initial chest CT studies and clinical data from 905 patients presenting between January 17 and March 3, 2020 to one of 18 centers in 13 provinces in China where patients had SARS-CoV-2 exposure, fever and a RT-PCR test for COVID-19. The exposure of SARS-CoV-2 is defined as either 1) travel history to Wuhan for patients collected outside of Wuhan, or travel to the animal market within 30 days of the symptom onset for patients who live in Wuhan or 2) close contact with patients with RTPCR confirmed SARS-CoV-2 infection for all patients. Of the 905 patients included in the study, 419 had a positive RT-PCR test while 486 had a negative test (confirmed by at least two additional negative RT-PCR tests and clinical observation).

### Clinical Information

Patient’s age, sex, exposure history, symptoms (present or absent of fever, cough and/or sputum), white blood cell counts, absolute neutrophil number, percentage neutrophils, absolute lymphocyte number and percentage lymphocytes were collected (Table 1). Sex, exposure history and symptoms were categorical variables. We used the LabelEncoder function in scikit-learn package to encode the target categorical variables into numerical variables. Then, we normalized each feature within a range of 0 and 1 using MinMaxScaler function in the scikit-learn package for further model development.

### Reader studies

The predictions of the AI models were compared to two radiologists on the test set. Both radiologists were board-certified (A.J. chest fellowship trained with 10 years’ clinical experience; S.S a current chest radiology fellow). The readers were given patients’ initial CT scan (at presentation) and associated clinical history that were used to test the AI models. Each reader independently reviewed the same set and evaluated the initial CT scan and clinical details, and combined imaging and clinical data in their review in a manner consistent with their clinical practice. Using this data, they predicted the COVID-19 status of the patients. Their predictions were compared to those of the AI algorithm and the RT-PCR results.

### AI Models

The RT-PCR virology test (COVID-19 (+) or COVID-19 (-)) was used as the reference to train the models. We developed and evaluated three different models using CT images and clinical information. Firstly, a deep learning model using a convolutional neural network (Model 1) was developed to only use CT images to predict COVID-19 status. Secondly, conventional machine learning methods (Model 2), including support vector machine (SVM), random forest and multi-layer perceptron (MLP), were evaluated to predict COVID-19 using only clinical information. Finally, we created a joint convolutional neural network model (Model 3) combining the radiologic data and the clinical data.

### Convolutional Neural Network Model (Model 1)

We proposed a CNN-based AI system to diagnose COVID-19 using a full CT scan of the chest. Similar to the previously reported AI diagnosis system^17,18^, our algorithm consisted of two CNN subsystems to firstly identify the abnormal CT slices and then to perform region-specific disease diagnosis (Fig. 1). More specifically, the slice selection CNN was trained to evaluate a chest CT slice and assign a probability that it was normal. The inverse of this probability was then used to rank the abnormal slices of each CT scan. We selected the 10 most abnormal slices from each study for the subsequent disease diagnosis due to the tradeoff of efficiency and turn-around time. The disease diagnosis CNN was designed to classify COVID-19 patients using multiple instance learning. The CNN was trained to predict whether a CT slice is from a COVID-19 (+) or COVID-19 (-) patient. The average probability from the 10 abnormal CT slices from each patient’s study was used to generate a prediction of COVID-19 status for the patient.

### Image Preprocessing

The first step is to select pertinent slices from the hundreds of images produced by a CT scan. Pertinent images contain pulmonary tissue and a potential parenchymal abnormality. For the selection of pertinent slices, image segmentation was used to detect parenchymal tissue. The raw CT images all had a 512 x 512 matrix storing CT intensities in Hounsfield Units (HU). A standard lung window (width (w)=1500 HU and level (l)=-600 HU) was used to normalize each slice to pixel intensities between 0 and 255. We segmented CT images into two parts, body and lung. The body part was segmented by finding the largest connected component consisting of pixels with an intensity greater than 175. The segmented connected component was filled into a solid region. The lung region was defined as the pixels with intensity less than 175 that fall within the segmented body part. Small regions with less than 64 pixels were removed, as they are typically segmented due to random noise. The lung region was enlarged by 10 pixels to fully include the pleural boundary. We discard images if the size of the lung was smaller than 20% of the size of the body part.

#### 1) Slice Selection CNN

We used a pre-trained Inception-ResNet-v2^19^ model based on the ImageNet^20^ as the slice selection CNN to identify abnormal CT images from all chest CT images^21^. The slice selection CNN was pre-trained in a previous pulmonary tuberculosis (PTB) detection study on CT images from a total of 484 non-TB pneumonia patients, including bacterial pneumonia, viral pneumonia and fungal pneumonia, in addition to 439 PTB and 155 normal chest CT patients. For CT images, the TB model predicts the probabilities of 3 classes, including pulmonary tuberculosis, non-TB pneumonia and normal chest CT. This model achieved 99.4% accuracy in differentiating normal slices from abnormal (PTB and non-TB pneumonia) slices. In this work, we applied the PTB model to a full CT scan to select 10 slices with the lowest probability of being normal. We noted that these selected slices may show no abnormal findings if the COVID-19 (+/-) patient’s CT is normal.

#### 2) Disease Diagnosis CNN

We used the 18-layer residual network (ResNet-18^22^) as the disease diagnosis CNN. The ResNet-18 takes images of segmented lungs as input and outputs probability of COVID-19 positivity. A max pooling layer that outputs log probability was used at the last layer, instead of the standard design that uses an average pooling layer at the last layer. The rationale of this design is that, given the abnormal finding is usually localized in a subregion of a CT image, we would like to predict whether a small region is abnormal due to COVID-19^23^. The CNN model can then be seen as a classifier that reports whether its receptive field is COVID-19 (+). The label of an image is then predicted by combining all predictions of every local region over the whole image. Max pooling serves as an “OR” gate that labels an image as COVID-19 (+) if there is any subregion in it that is COVID-19 (+). Patient level prediction was set as the average of image-level prediction of a patient’s 10 most abnormal images. To visualize the CNN’s prediction, we up-sampled the CNN’s outputs, without applying the max pooling layer, to the original image size. The lung mask was applied to up-sampled outputs for clear visualization.

#### 3) CNN Training

We used binary cross entropy as the objective function. Adam optimizer^24^ with a learning rate 0.001 was used to train the neural network. The learning rate was decreased by a factor of 0.95 each epoch. We applied random rotation, grid distortion, and cutout^25^ to images for data augmentation. 20% of training samples were held out as the tuning set to monitor the progress of the training process. The training process was iterated for 40 epochs with a batch size of 16 samples. Performance on the tuning set was monitored every 100 iterations. The model with lowest binary cross entropy on the tuning set was selected as the final model. Parameters were tuned to ensure that the validation error decreases along with the training error.

We designed a weakly supervised task to initialize weights of the CNN model. Specifically, we randomly selected image patches of lung regions from training images and labeled those patches as the label of the training images. The CNN is then pre-trained to classify these image patches for 1 epoch. This weakly supervised task accords with the idea that the CNN is classifying a local region in the CT image to be COVID-19 (+/-).

### Machine Learning Classifiers (Model 2)

We developed support vector machine, random forest and multi-layer perceptron classifiers based on patients’ age, sex, exposure history, symptoms (present or absent of fever, cough and/or sputum), white blood cell counts, neutrophil counts, percentage neutrophils, lymphocyte counts and percentage lymphocytes. We fine-tuned the hyperparameters of each classifier on the training set and tuning set, and evaluated the best model on the test set. For the support vector machine classifier, we assessed the “C”, and kernel. For the random forest classifier, the number of estimators was tuned. For multi-layer perceptron, we assessed the number of layers and the number of hidden nodes in each layer. After the hyperparameter optimization, a 3-layer MLP model with 64 nodes in each layer was selected because of the highest AUC score on the tuning set The MLP model was selected because of the highest AUC score on the tuning set (Extended Data Fig. 4). We used the Scikit-learn^26^ package to fit and evaluate these models.

### Joint Model: Combining CT imaging and clinical information (Model 3)

We trained a model to integrate CT imaging data and clinical information. We applied the global averaging layer to the last layers of the convolutional model described previously to derive a 512 dimensional feature vector to represent a CT image. A total of 12 clinical features (Table 1) of the same patient were concatenated with this feature vector. A MLP takes this combined feature vector as the input to predict the status of COVID-19. We used a 3-layer MLP, each layer has 64 nodes and is composed by a batch normalization layer, a fully connected layer and a ReLU activation function. Normalized Gaussian noise was added at the input layers for data augmentation. The MLP was jointly trained with the CNN. We applied binary cross entropy to validate the predictions from both MLP and CNN during the training process. The sum of these two measurements was used as the overall objective function to train the joint model. We used the same optimization strategy of Model 1 to train the MLP and CNN, except that the learning rate was increased to 0.002. The CNN was also initialized by the weakly supervised task of classifying the small image patches.

### Statistical analysis

The 95% confidence intervals of sensitivity, specificity and accuracy were calculated by the exact Clopper-Pearson method^27^. The confidence intervals of AUC were calculated by the DeLong methods^28^. The confidence interval of predictive values was calculated by the standard logit confidence intervals^29^. McNemar’s test^30^ was used to calculate the two-sided p value for sensitivity and specificity between models and human readers. The Youden index was used to determine the optimal model sensitivity and specificity. Statistical significance was defined as a p-value less than 0.05. Logistic regression was used to evaluate the significance of each clinical variable. Hosmer-Lemeshow goodness of fit^31^ was used to assess the goodness of fit of the logistic regression. The statistics of AUC comparisons were computed in the *pROC* package^32^. McNemar’s test and the exact confidence intervals were calculated in the statsmodels package in python.

### Reporting Summary

Further information on research design is available in the Nature Research Reporting Summary linked to this article.

### Code Availability

The code used for training the deep learning models and the pre-trained models and models used in this study are all available at https://github.com/howchihlee/COVID19_CT. Implementations of our work is based on following open source repositories: Tensorflow: https://www.tensorflow.org; Pytorch: https://pytorch.org; Keras: https://keras.io; Sklearn: https://scikit-learn.org/stable/. pROC: https://cran.r-project.org/web/packages/pROC/index.html. statsmodels: https://www.statsmodels.org/stable/index.html.

---

**Extended Data Fig. 1.**
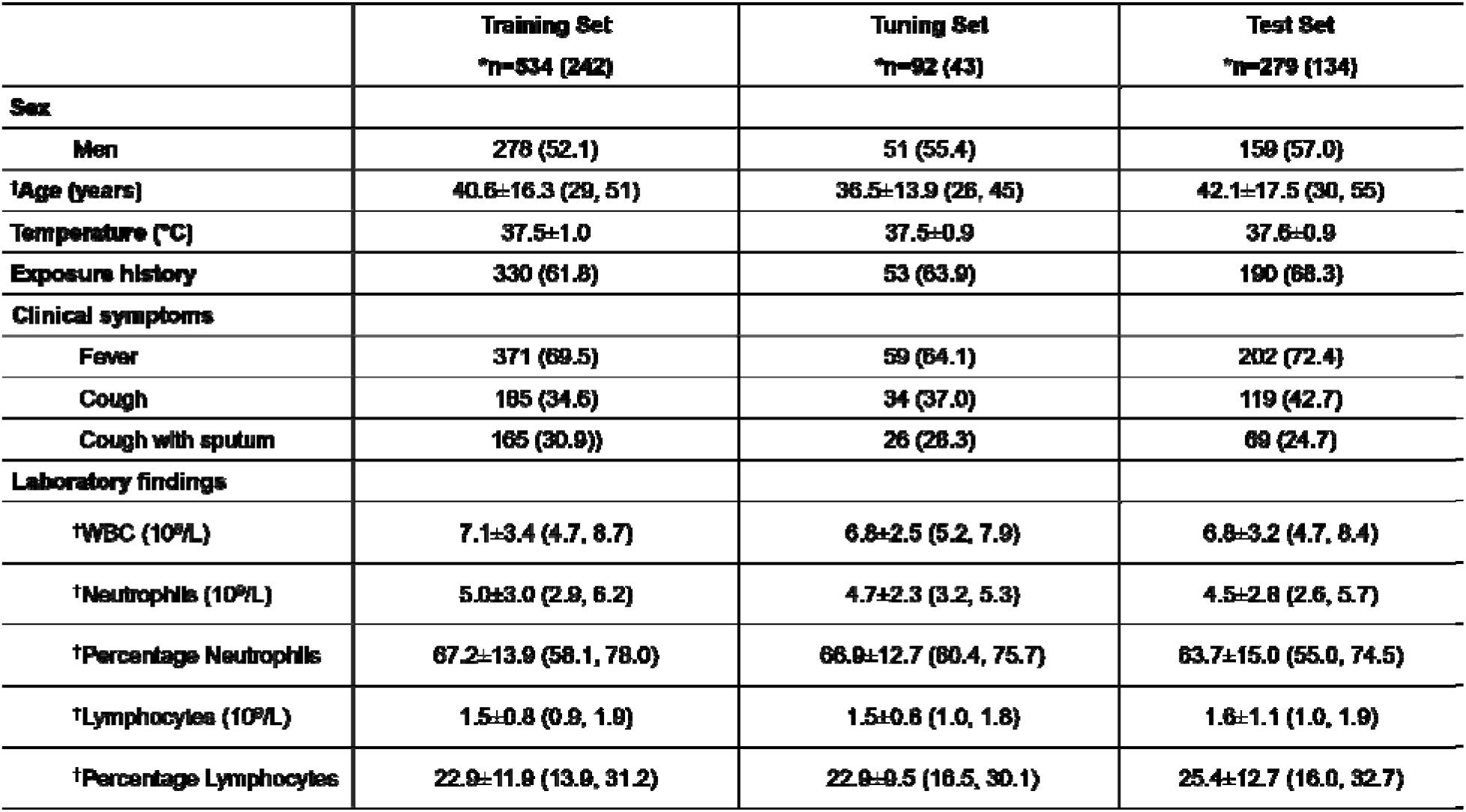
Characteristics of clinical features in the training set, tuning set and test set. ± indicates mean ± standard deviation * Data in parenthesis shows the number of COVID-19 positive patients † Data in parenthesis shows Interquartile Range (IQR) Other data in parenthesis shows percentage

**Extended Data Fig. 2.**
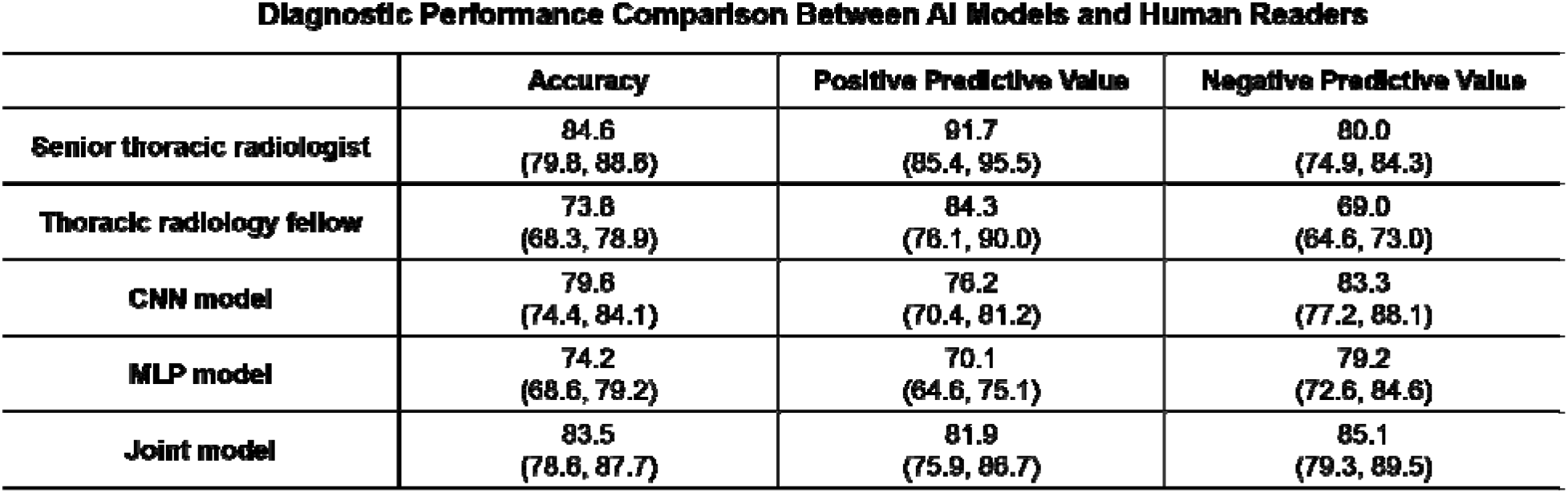
Comparisons of the diagnostic performance between AI models and human readers on a test set of 279 cases. Data were presented in percentage and the 95% confidence interval. The confidence intervals of accuracy were calculated by the exact Clopper-Pearson method. The confidence intervals of the predictive values were calculated by the standard logit confidence intervals.

**Extended Data Fig. 3.**
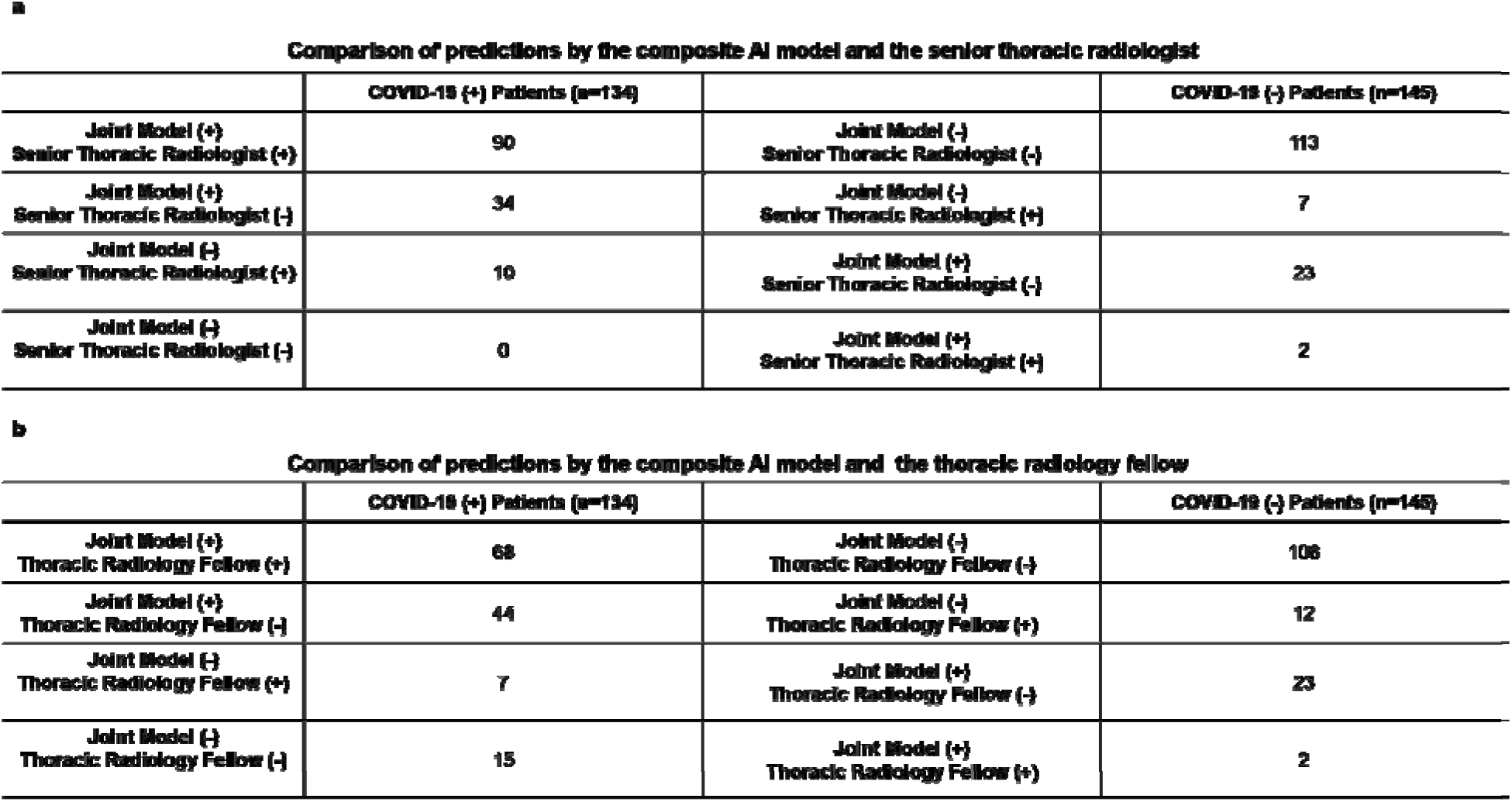
Comparisons of predictions by the joint model and human readers on a test set of 279 cases. The (+/-) indicates the prediction of the COVID-19 status by the joint model and human readers.

**Extended Data Fig. 4.**
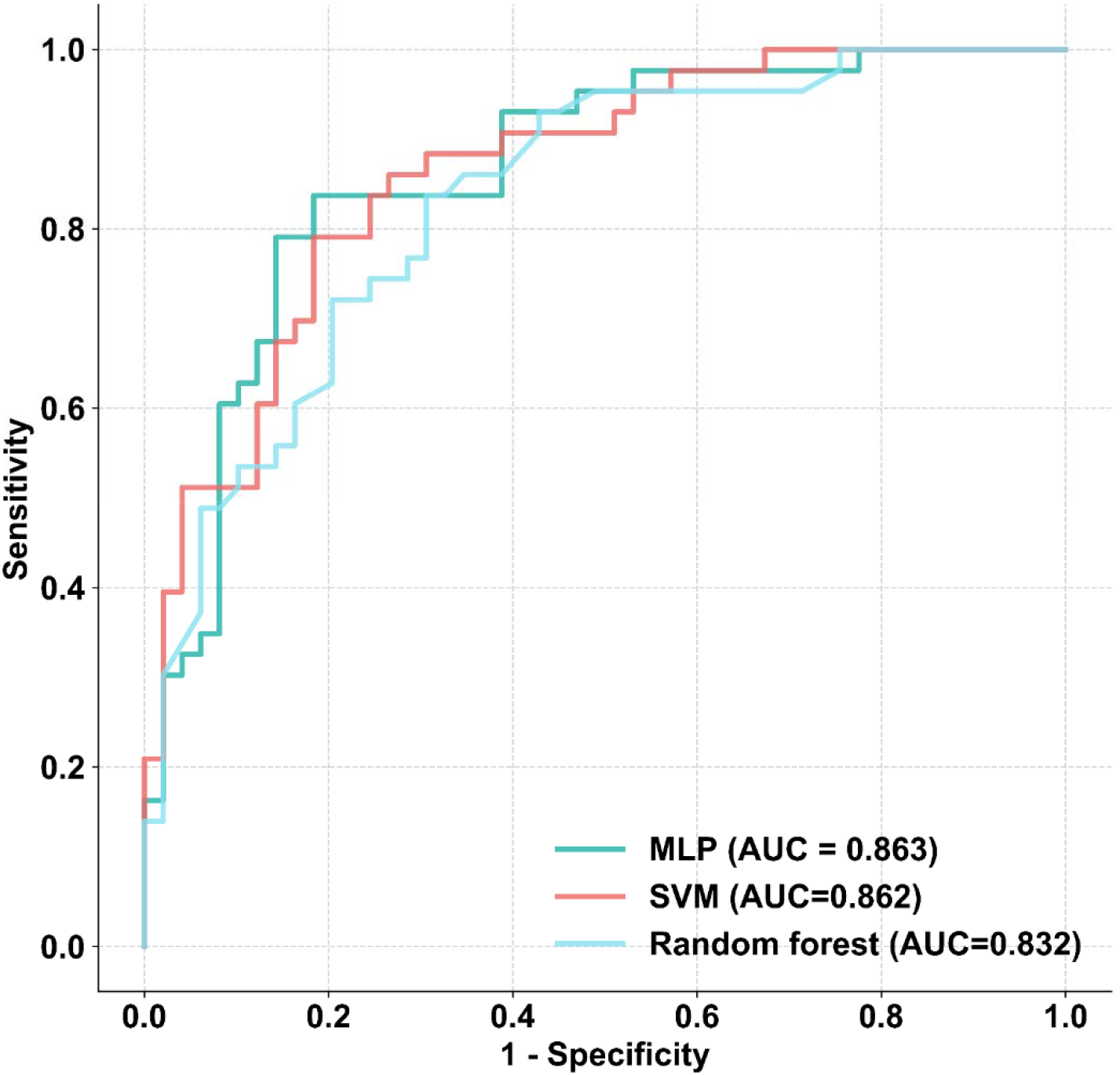
ROC curve comparison of the MLP, random forest and SVM models on the tuning set of 92 cases. Two-sided p-value indicates the significance of difference in performance metric compared with respect to the MLP model by using the DeLong test. The MLP showed no significant difference as compared to the SVM model (*p*=0.98) and the random forest model (*p*=0.35). The MLP model was selected in this study due to the highest AUC score.

## References

1. Zhu, N., et al. A novel coronavirus from patients with pneumonia in China, 2019. N Engl J Med 382, 727–733 (2020).

2. Tan, W., et al. A novel coronavirus genome identified in a cluster of pneumonia cases—Wuhan, China 2019− 2020. China CDC Weekly 2, 61–62 (2020).

3. Chan, J.F., et al. A familial cluster of pneumonia associated with the 2019 novel coronavirus indicating person-to-person transmission: a study of a family cluster. Lancet 395, 514–523 (2020).

4. Huang, C., et al. Clinical features of patients infected with 2019 novel coronavirus in Wuhan, China. Lancet 395, 497–506 (2020).

5. Phan, L.T., et al. Importation and Human-to-Human Transmission of a Novel Coronavirus in Vietnam. N Engl J Med 382, 872–874 (2020).

6. Li, Q., et al. Early Transmission Dynamics in Wuhan, China, of Novel Coronavirus-Infected Pneumonia. N Engl J Med 382, 1199–1207 (2020).

7. Chung, M., et al. CT Imaging Features of 2019 Novel Coronavirus (2019-nCoV). Radiology 295, 202–207 (2020).

8. Phelan, A.L., Katz, R. & Gostin, L.O. The Novel Coronavirus Originating in Wuhan, China: Challenges for Global Health Governance. JAMA 323, 709–710 (2020).

9. Nishiura, H., et al. The Extent of Transmission of Novel Coronavirus in Wuhan, China, 2020. J Clin Med 9, 330 (2020).

10. Xie, X., et al. Chest CT for Typical 2019-nCoV Pneumonia: Relationship to Negative RT-PCR Testing. Radiology, https://doi.org/10.1148/radiol.2020200343 (2020).

11. Wang, D., et al. Clinical Characteristics of 138 Hospitalized Patients With 2019 Novel Coronavirus-Infected Pneumonia in Wuhan, China. JAMA 323, 1061–1069 (2020).

12. Kanne, J.P. Chest CT Findings in 2019 Novel Coronavirus (2019-nCoV) Infections from Wuhan, China: Key Points for the Radiologist. Radiology 295, 16–17 (2020).

13. Song, F., et al. Emerging 2019 Novel Coronavirus (2019-nCoV) Pneumonia. Radiology 295, 210–217 (2020).

14. World Health Organization. Clinical management of severe acute respiratory infection when novel coronavirus (2019-nCoV) infection is suspected: interim guidance, 28 January 2020. (WHO, 2020).

15. Ai, T., et al. Correlation of Chest CT and RT-PCR Testing in Coronavirus Disease 2019 (COVID-19) in China: A Report of 1014 Cases. Radiology, https://doi.org/10.1148/radiol.2020200642 (2020).

16. Li, L., et al. Artificial intelligence distinguishes covid-19 from community acquired pneumonia on chest ct. Radiology, https://doi.org/10.1148/radiol.2020200905 (2020).

## Methods-only References

17. Liu, F., et al. Deep Learning Approach for Evaluating Knee MR Images: Achieving High Diagnostic Performance for Cartilage Lesion Detection. Radiology 289, 160–169 (2018).

18. Liu, F., et al. Fully Automated Diagnosis of Anterior Cruciate Ligament Tears on Knee MR Images by Using Deep Learning. Radiol Artif Intell 1, 180091 (2019).

19. Szegedy, C., Ioffe, S., Vanhoucke, V. & Alemi, A.A. Inception-v4, inception-resnet and the impact of residual connections on learning. in Thirty-first AAAI conference on artificial intelligence Preprint at https://arxiv.org/abs/1602.07261 (2017).

20. Russakovsky, O., et al. Imagenet large scale visual recognition challenge. Intl J Comput Vis 115, 211–252 (2015).

21. Wang, Y., et al. A Generalized Deep Learning Approach for Evaluating Secondary Pulmonary Tuberculosis on Chest Computed Tomography. Preprint with the Lancet (2019).

22. He, K., Zhang, X., Ren, S. & Sun, J. Deep residual learning for image recognition. in Proceedings of the IEEE conference on computer vision and pattern recognition 770–778 (2016).

23. Oquab, M., Bottou, L., Laptev, I. & Sivic, J. Is object localization for free?-weakly-supervised learning with convolutional neural networks. in Proceedings of the IEEE conference on computer vision and pattern recognition 685–694 (2015).

24. Kingma, D.P. & Ba, J. Adam: a method for stochastic optimization. Preprint at https://arxiv.org/abs/1412.6980 (2014).

25. DeVries, T. & Taylor, G.W. Improved regularization of convolutional neural networks with cutout. Preprint at https://arxiv.org/abs/1708.04552 (2017).

26. Pedregosa, F., et al. Scikit-learn: Machine learning in Python. J Mach Learn Res 12, 2825–2830 (2011).

27. Agresti, A. & Coull, B.A. Approximate is better than “exact” for interval estimation of binomial proportions. Am Stat 52, 119–126 (1998).

28. DeLong, E.R., DeLong, D.M. & Clarke-Pearson, D.L. Comparing the areas under two or more correlated receiver operating characteristic curves: a nonparametric approach. Biometrics, 837–845 (1988).

29. Mercaldo, N.D., Lau, K.F. & Zhou, X.H. Confidence intervals for predictive values with an emphasis to case-control studies. Stat Med 26, 2170–2183 (2007).

30. McNemar, Q. Psychological statistics (Wiley, New York, 1962).

31. Hosmer, D.W., Hosmer, T., Le Cessie, S. & Lemeshow, S. A comparison of goodness of fit tests for the logistic regression model. Stat Med 16, 965–980 (1997).

32. Robin, X., et al. pROC: an open-source package for R and S+ to analyze and compare ROC curves. BMC Bioinform 12, 77 (2011).

